# Pharyngeal carriage of inoculated recombinant commensal bacteria generates antigen-specific immunological memory

**DOI:** 10.1101/2020.05.06.20093450

**Authors:** Jay R Laver, Diane Gbesemete, Adam P Dale, Zoe C Pounce, Carl N Webb, Eleanor F Roche, Graham Berreen, Konstantinos Belogiannis, Alison R Hill, Muktar M Ibrahim, David W Cleary, Anish K Pandey, Holly E Humphries, Lauren Allen, Hans de Graaf, Martin C Maiden, Saul N Faust, Andrew R Gorringe, Robert C Read

**Affiliations:** Faculty of Medicine and Institute for Life Sciences, University of Southampton, Southampton, UK; NIHR Southampton Biomedical Research Centre and NIHR Southampton Clinical Research Facility, University Hospital Southampton NHS Foundation Trust, Southampton, UK; Public Health England, Porton Down, Salisbury, UK; Department of Zoology, University of Oxford, Oxford, UK

## Abstract

The human nasopharynx is colonized by commensal bacteria and pathobionts, which comprise a complex microbial ecosystem capable of generating primary and secondary immune responses. Experimental intranasal infection of human adults with the commensal *Neisseria lactamica* results in safe, sustained colonization. Herein is described a novel technology to chromosomally transform *N. lactamica* with heterologous antigen, for the purpose of safe delivery to the mucosal surface and the generation of an antigen-specific immune response. *N. lactamica* was transformed to express the meningococcal vaccine antigen Neisseria Adhesin A (NadA) and was inoculated intranasally into humans at a dose of 10^5^ colony-forming units. NadA-expressing *N. lactamica* colonized these individuals and was carried asymptomatically for 3 months. Colonization with NadA-expressing *N. lactamica* generated NadA-specific IgG-secreting plasma cells within 14 days of colonization and both NadA-specific IgG and NadA-specific IgG memory B cells within 28 days of colonization. NadA-specific IgG memory B cells circulate in the bloodstream of colonized participants for at least 90 days. Genetically transformed *N. lactamica* has the potential to be a safe bacterial vehicle to generate beneficial immune responses to a wide range of heterologous antigens during sustained pharyngeal carriage.

## MAIN

Natural protective immunity to invasive disease caused by nasopharyngeal pathobionts including *Streptococcus pneumoniae, Haemophilus influenzae* and *Neisseria meningitidis* (Nmen) is a consequence of repetitive, transient, asymptomatic carriage commencing in infancy. In each case, carriage is associated with seroconversion against cognate antigen^1-3^. This natural mechanism could be harnessed using safe genetically transformed live bacterial vectors. The human commensal bacterium *Neisseria lactamica* (Nlac) is a harmless colonizer of babies and young children. The frequency of colonization wanes during early childhood with niche replacement by the closely related pathobiont Nmen^4^. Controlled human intranasal infection of wild type (WT) Nlac results in safe colonization of human volunteers, which is sustained for at least 6 months, and is accompanied by humoral and mucosal immune responses^5^, and reduced natural acquisition of Nmen^6^. During experimental human colonization the genome of Nlac remains stable^7^. This, together with the inherent adjuvant properties of its outer membrane components^8^ suggests the bacterium could be adapted as a microbial factory/delivery platform, producing molecules of biological or therapeutic relevance, such as vaccine antigens, *in situ* following colonization^9-11^. Nlac is relatively resistant to genetic manipulation^12^ due to its repertoire of restriction endonucleases. Therefore we developed a novel cloning system to integrate heterologous genes into the chromosome using hypermethylated nucleic acids (Supplementary Figure 1). The ability to transform Nlac confirmed it is naturally competent (Supplementary Figure 2), with horizontal gene transfer affected by similar factors to other members of the genus (Supplementary Figure 3). The culmination of these discoveries is a means to chromosomally transform gene constructs into the Nlac chromosome whilst avoiding the use of antibiotic resistance markers, which is critical to prevent the unnecessary spread of antimicrobial resistances. The methodology exploits the natural ability of Nlac to ferment lactose via the activity of the enzyme β-galactosidase (coded for by the *lacZ* gene) and utilises blue/white screening on media containing 5-bromo-4-chloro-3-indolyl-β-D-galactopyranoside (X-gal) to screen for mutants (Supplementary Figure 4). High-level expression of heterologous genes is achieved using a novel, synthetic promoter that exploits the transcription enhancement property of DNA preceding the meningococcal *porA* gene (Supplementary Figure 5).

The genomically-defined Nlac strain Y92-1009^13^ was transformed with a construct containing the coding sequence for *Neisseria* adhesin A (NadA), a meningococcal-specific member of the type V autotransporter family of outer membrane proteins^14^ to generate strain 4NB1. NadA is one of the strongly immunogenic, recombinant protein components of the 4CMenB vaccine (Bexsero), and the only one capable of generating sterilizing immunity following immunisation of a transgenic mouse model of meningococcal colonization^15^. NadA expression in Nmen is associated with an increased level of adhesion to human epithelial cell lines^14^, but is expressed in a minority of Nmen lineages so is non-essential for virulence^16^. A second (control) strain (4YB2) was transformed with a construct otherwise identical to the first, except for the replacement of the *nadA* coding sequence with a 14 bp, non-coding linker sequence (Supplementary Figure 6).

These recombinant strains were used to conduct a first-in-man, controlled human infection model experiment (CHIME), with the co-primary objectives:

1. To establish the safety of nasal inoculation of healthy volunteers with a genetically modified strain of *Neisseria lactamica* expressing NadA.
2. To assess the NadA-specific immunity in healthy volunteers following nasal inoculation with *Neisseria lactamica* expressing NadA.

The clinical protocol for the CHIME has been published elsewhere^17^. Deliberate Release of these genetically modified organisms in a CHIME was approved by the UK government Department of Farming and Rural Affairs (https://www.gov.uk/government/publications/genetically-modified-organisms-university-of-southampton-17r5001).

## RESULTS

### Expression of *nadA* in Nlac results in surface-expressed, immunogenic NadA and increased adherence to epithelial cells

The expression of *nadA* in the meningococcus is phase variable^18^ and regulated by the NadR repressor protein, which becomes de-repressed by salivary concentrations of the metabolite 4-hydroxyphenylacetic acid (4HPA)^19^. A longitudinal analysis of serial Nmen isolates recovered from the human nasopharynx shows that NadA expression decreases over time, likely a result of seroconversion against NadA and antibody-mediated selective pressure against NadA expression^20^. To prevent the gene from becoming phase ‘OFF’ in strain 4NB1 during the CHIME, transcription of the *nadA* gene in our construct is driven by a hybrid, non-phase variable *porA/porB* promoter. The transcription activity of this hybrid promoter is enhanced by 200 nucleotides of ‘upstream activation sequence’ (UAS) of the WT *porA* promoter, which is optimal for increasing gene expression in our constructs (Supplementary Figure 5).

Flow cytometry using anti-NadA monoclonal antibody (mAb) 6e3 (Figure 1A) demonstrates that Nlac strain 4NB1 expresses NadA on its surface, in contrast to both WT Y92-1009 and control strain 4YB2. Using an anti-NadA polyclonal antibody (pAb), NadA expression on the surface of strain 4NB1 is comparable to that of Nmen strain N54.1, a serogroup Y WT carriage isolate shown to abundantly express NadA, in comparison to other laboratory strains of the meningococcus^21^. This suggests the abundance of NadA protein on strain 4NB1 is equivalent to that of naturally circulating, NadA-expressing meningococci (Figure 1B). Compared to WT Y92-1009, strain 4NB1 shows increased association with confluent monolayers of HEp-2 epithelial cells (Figure 1C), consistent with the published role of NadA as an outer membrane adhesin^14^.

**Figure 1:**
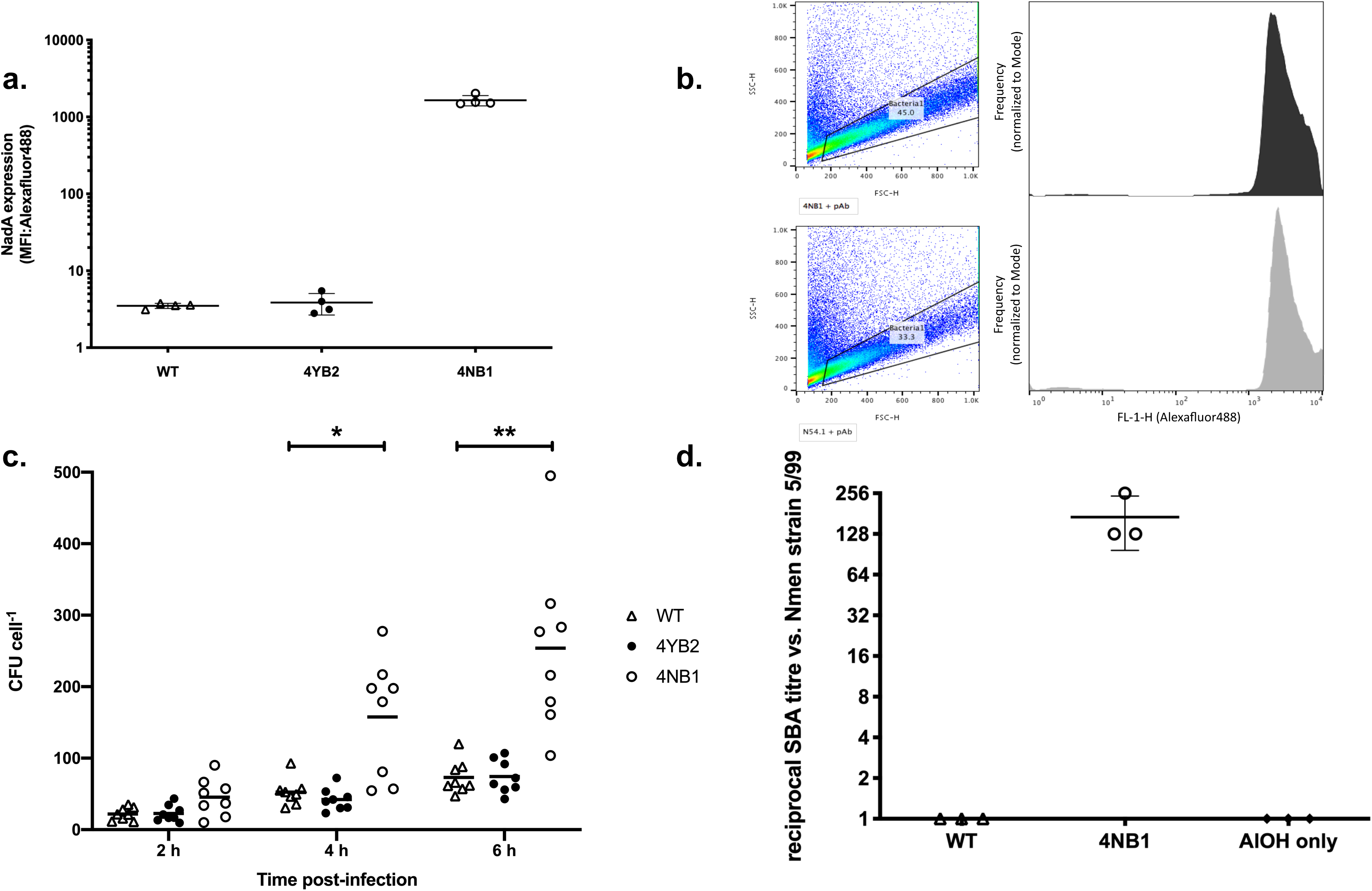
*N. lactamica-expressed* NadA functions as an adhesin and is immunogenic in mice. (**a**) MFI of cultures of WT Nlac strain Y92-1009 (∆) and the GM derivatives 4YB2 (•) and 4NB1 (○), labeled with murine anti-NadA monoclonal antibody 6e3 and goat-derived, anti-mouse IgG conjugated to Alexafluor488. Bars represent Mean ± SD, n = 4. (**b**) Gating strategy and fluorescence histograms (FL-1-H: Alexafluor488) of NadA-expressing WT Nmen N54.1 (*bottom, grey*) and GM-Nlac 4NB1 (*top, black*), each grown to mid-log phase in the presence of 5 mM 4HPA and then labeled with murine anti-NadA polyclonal serum and goat-derived, anti-mouse IgG conjugated to Alexafluor488. FSC-H: forward scatter, SSC-H: side scatter. Frequency of histogram is normalised to Mode. (**c**) Association of WT Nlac Y92-1009 (∆) and the GM derivatives 4YB2 (•) and 4NB1 (○) with monolayers of HEP-2 epithelial cells over time, presented as colony forming units (CFU) per cell. Bars indicate Mean. \**p* ≤ 0.05, \*\**p* ≤ 0.01 2-way ANOVA with Dunnett’s multiple comparisons test vs. Mean of ‘WT’ at each time point (n = 8). (**d**) Reciprocal endpoint serum bactericidal antibody titer measured in triplicate assays of pooled mouse sera vs. the NadA-overexpressing Nmen 5/99. Mice (n = 10 per group) were immunized intraperitoneally with either: (i) dOMV derived from either WT Nlac Y92-1009 (∆), or the GM derivative 4NB1 (○) supplemented with aluminium hydroxide (AIOH) as adjuvant, or (ii) AIOH alone (u). Bars represent Mean ± SD.

Immunisation of mice with deoxycholate-extracted outer membrane vesicles (dOMV) derived from strain 4NB1 (4NB1-dOMV), generated potent serum bactericidal antibody (SBA) activity against Nmen strain 5/99, a NadA-overexpressing reference strain^22^. In contrast, immunising mice with WT Y92-1009 dOMV generates no SBA (Figure 1D). This is consistent with previous observations in humans that antibody responses directed against the native outer membrane proteins of Nlac are not bactericidal^23^, and suggests that the SBA activity is instead directed through antibody targeting NadA.

### Pathogenic potential of recombinant Nlac is equivalent to wild type Nlac

To gain approval for Deliberate Release of GM-Nlac in a CHIME, it was necessary to demonstrate absence of pathogenic potential. Nmen expresses polysaccharide capsule^24^, which affords resistance to killing by human serum and constitutes the key determinant of the organism’s ability to cause invasive disease. It is plausible that Nlac might become more pathogenic if it could deposit polysaccharide capsule on its surface. Given the propensity of Nlac to take up DNA containing *Neisseria* DNA uptake sequences (DUS) (see Supplementary Information), the most likely potential source of ectopic capsule synthesis genes during human colonization is Nmen, in which the *Neisseria* DUS is overrepresented^25^, and which shares the nasopharyngeal niche with Nlac. The uptake and assimilation of an antibiotic resistance marker (gene *aphA3*, coding Kan^R^) from three different genomic sources into the chromosome of WT Nlac strain Y92-1009, GM-Nlac strain 4NB1 and Nmen strain MC58 was therefore measured (Figure 2A). Genomic DNA (gDNA) derived from Y92-1009*∆nlaIII:aphA3* efficiently transformed both WT Y92-1009 and strain 4NB1, consistent with donor and recipient strains having homologous patterns of DNA methylation, which precludes degradation of the gDNA by restriction endonuclease activities and increases transformation efficiency (TE). The same gDNA also transformed Nmen strain MC58, although less efficiently. Conversely, whilst gDNA derived from mutants MC58*∆siaD:aphA3* (in which the Kan^R^ gene inactivates a capsule biosynthesis gene) and MC58*∆nadA:aphA3* (in which the Kan^R^ gene inactivates the endogenous *nadA* gene) was able to transform MC58; neither gDNA was able to generate a single Nlac transformant (n = 5 experiments). The TE of 4NB1 with Nmen gDNA, under conditions ideal for transformation, is therefore < 1 transformant per 2 × 10^9^ CFU, and is the same as the TE of the parental strain. We conclude that the likelihood of GM-Nlac assimilating capsule synthesis genes from Nmen remains as low as the WT parental strain and that the risk of this occurring *in vivo* is negligible.

**Figure 2:**
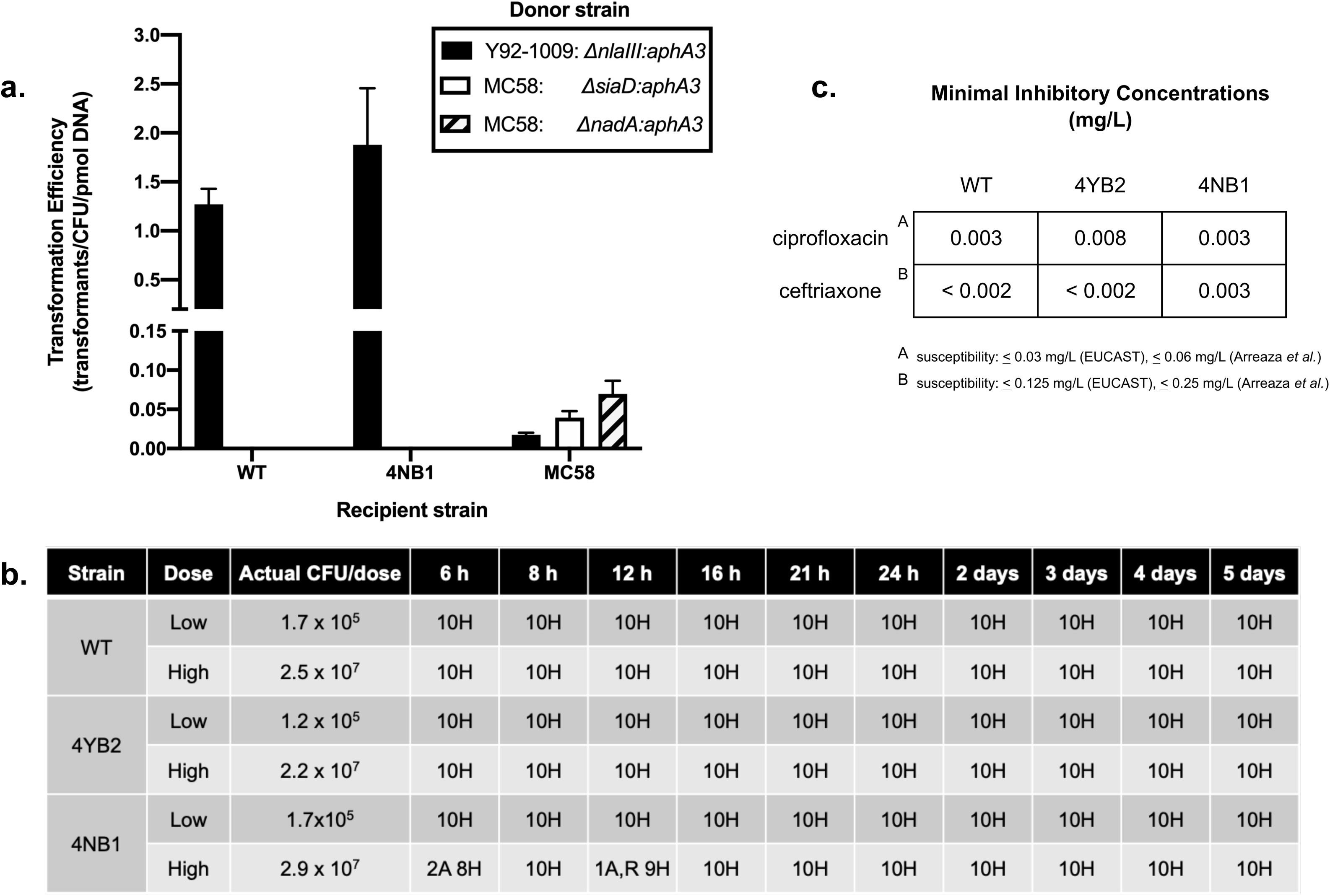
Genetically modified *N. lactamica* is non-pathogenic in a murine model of systemic infection. (**a**) Transformation efficiency (TE) of WT Nlac Y92-1009, the NadA-expressing GM-Nlac strain 4NB1 and WT Nmen strain MC58 with 0.0001 pmol of gDNA from one of three *Neisseria* strains: (i) Nlac Y92-1009*AnlaIII:aphA3 (black bars)*, (ii) Nmen MC58*∆siaD:aphA3 (white bars)* and (iii) Nmen MC58*∆nadA:aphA3 (hatched bars)*. Bars represent Mean ± SD (n = 5). (**b**) Qualitative health scores for groups of mice inoculated intraperitoneally with ‘high’ and ‘low’ doses of one of WT Nlac Y92-1009, or the GM-Nlac strains 4YB2 and 4NB1. Bacteria were injected, along with 10 mg human holo-transferrin on Day 0. A second injection of 10 mg human holo-transferrin was made after 24 h. Mice were regularly monitored over 5 days. The number of mice assessed as exhibiting normal or ‘healthy’ (H) behaviour, and the number of mice exhibiting classic signs of mild discomfort such as having an ‘arched’ (A) or ‘ruffled’ (R) appearance were recorded. After the first day no mice exhibited any signs of discomfort. (**c**) MICs (mg L^-1^) of antibiotics used clinically to treat meningococcal disease versus GM-Nlac strain 4NB1, as determined by e-test. Tests carried out according to European Committee on Antimicrobial Susceptibility Testing (EUCAST) methodology.

Neither strain 4NB1 nor 4YB2 proved any more pathogenic than WT Y92-1009 in a mouse infection model at two different inoculum doses (Figure 2B). Both GM-Nlac strains retained susceptibility to standard antibiotics for the treatment of meningococcal disease (Figure 2C). E-tests demonstrated that the minimal inhibitory concentrations (MICs) of ciprofloxacin and ceftriaxone for GM-Nlac were well below the European Committee on Antimicrobial Susceptibility Testing (EUCAST) breakpoints for Nmen and those previously established in 286 Nlac isolates^26^.

### GM-Nlac safely colonizes the human upper respiratory tract in a CHIME

Amongst 42 volunteers screened, 26 were free from Nlac and Nmen carriage and eligible for participation (Figure 3A). Participants were assigned, initially in 1-person but subsequently in up-to-5-person blocks, to either the ‘intervention’ (strain 4NB1) or ‘control’ (strain 4YB2) study arm. Volunteers were admitted for observation to the NIHR Clinical Research Facility (CRF) at University Hospital Southampton for the first 4.5 days after inoculation (i.e. the ‘admission period’). Volunteers were deemed ‘colonized’ by GM-Nlac following recovery of one or more colonies of GM-Nlac from either nasal wash fluid or throat swabs at any point within 14 days of inoculation. Eligible volunteers were inoculated in each study block until at least 11 volunteers had been successfully colonized. The colonization status of Nlac and Nmen for each participant during the follow-up period over 90 days is shown in Figure 3B.

**Figure 3:**
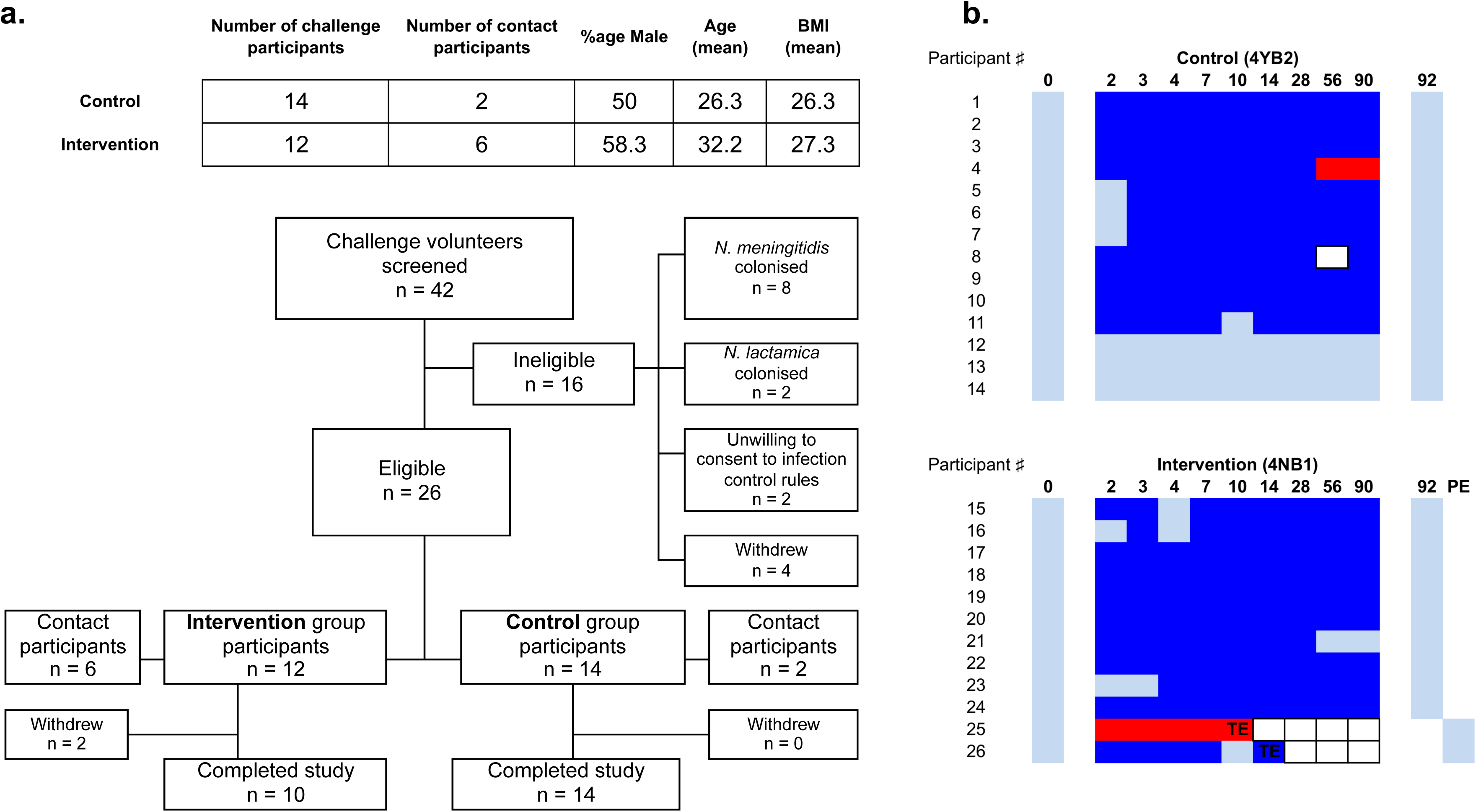
Controlled human infection model experiment using genetically modified *N. lactamica*. (**a**) *(Top)* Demographic data of participants challenged with the control, WT-equivalent strain of GM-Nlac (4YB2, **Control**) or the NadA-expressing strain of GM-Nlac (4NB1, **Intervention**). (*Bottom*) Study flow diagram showing allocation to groups and study completion. (**b**) Pattern of *Neisseria* species carriage for all volunteers throughout the course of the study, showing **Control** and **Intervention** groups. Participant numbers are assigned arbitrarily. Days post-inoculation at time of sampling are shown in bold. Color blocks represent: nonattendance (*white, bordered*), no *Neisseria* species cultured (*light blue*), 1+ colonies of PCR-verified GM-Nlac cultured (*dark blue)*, Nmen cultured in the absence of Nlac/GM-Nlac (*red*). Abbreviations: **TE** – triggered eradication following withdrawal from study, **PE** – post-eradication visit to certify clearance of GM-Nlac (= **TE** + 2 days).

In the event of withdrawal from the study (n = 2) or at day 90 post-inoculation (n = 24), single dose oral ciprofloxacin (500 mg) was administered, which eliminated carriage of both strains of GM-Nlac within 2 days (Figure 3B). During both the admission period (Supplementary Figure 7) and the follow-up period (Supplementary Figure 8), there were only a small number of solicited and/or unsolicited symptoms reported, and similarly few clinically observed (Supplementary Figure 9) or laboratory test-identified (Supplementary Figure 10) adverse events recorded. Upon clinical review, none were deemed to be related or likely related to carriage of either GM-Nlac strain and there was no increase in adverse events in the intervention study arm in comparison to the control study arm. There were no serious adverse events, no antibiotic eradication or treatment given due to adverse events, and no study withdrawal due to adverse events. These data indicate that GM-Nlac colonization, as with the WT strain, is safe and asymptomatic. At no point throughout the study was GM-Nlac detected in exhaled breath samples or on surgical facemasks worn prior to each visit to the CRF (data not shown). Additionally, there was no detectable transmission of GM-Nlac to ‘bedroom-sharing contacts’ of volunteers, as established by culture of oropharyngeal swab taken from these individuals at multiple time points (n = 8, data not shown).

### Colonization with NadA-expressing GM-Nlac expands circulating Nlac-specific and NadA-specific, IgG-secreting cells

An enzyme-linked immunospot (ELISpot) assay was employed to detect IgG-secreting cells with specificity to one of the following: (i) dOMV derived from strain 4YB2 (4YB2-dOMV), (ii) 4NB1-dOMV, (iii) the soluble domain of NadA (sNadA). We compared the maximum (‘peak’) number of IgG-secreting spot-forming units (SFU) per 200,000 peripheral blood mononuclear cells (PBMC) developed against each antigen post-inoculation. Consistent with the rapid appearance and transient nature of circulating plasmablasts following infection or vaccination^27^, ‘peak’ responses were always measured in samples taken at either day 7 or day 14 post-inoculation (+7-14) (data not shown).

In 4YB2-colonized participants there was a significant increase in the number of SFU with specificity to either 4YB2-dOMV (Figure 4A) or 4NB1-dOMV (Figure 4B) in comparison to baseline, but no significant change in the number of SFU with specificity to sNadA (sNadA-specific SFU) (Figure 4C). Conversely, in 4NB1-colonized participants the peak number of sNadA-specific SFU was significantly increased over baseline (Figure 4F). Whilst the number of 4NB1-dOMV-specific SFU was significantly increased in these participants compared to baseline (Figure 4E), there was no difference between the baseline and peak measurements of 4YB2-dOMV-specific SFU (Figure 4D). This suggests that NadA is an immunodominant antigen in the context of expression on Nlac. The numbers of tetanus toxoid (TT)-specific SFU generated by colonized individuals at baseline and at intervals during colonization were unchanged, indicating an absence of non-specific ‘bystander’ antibody-secreting cells.

**Figure 4:**
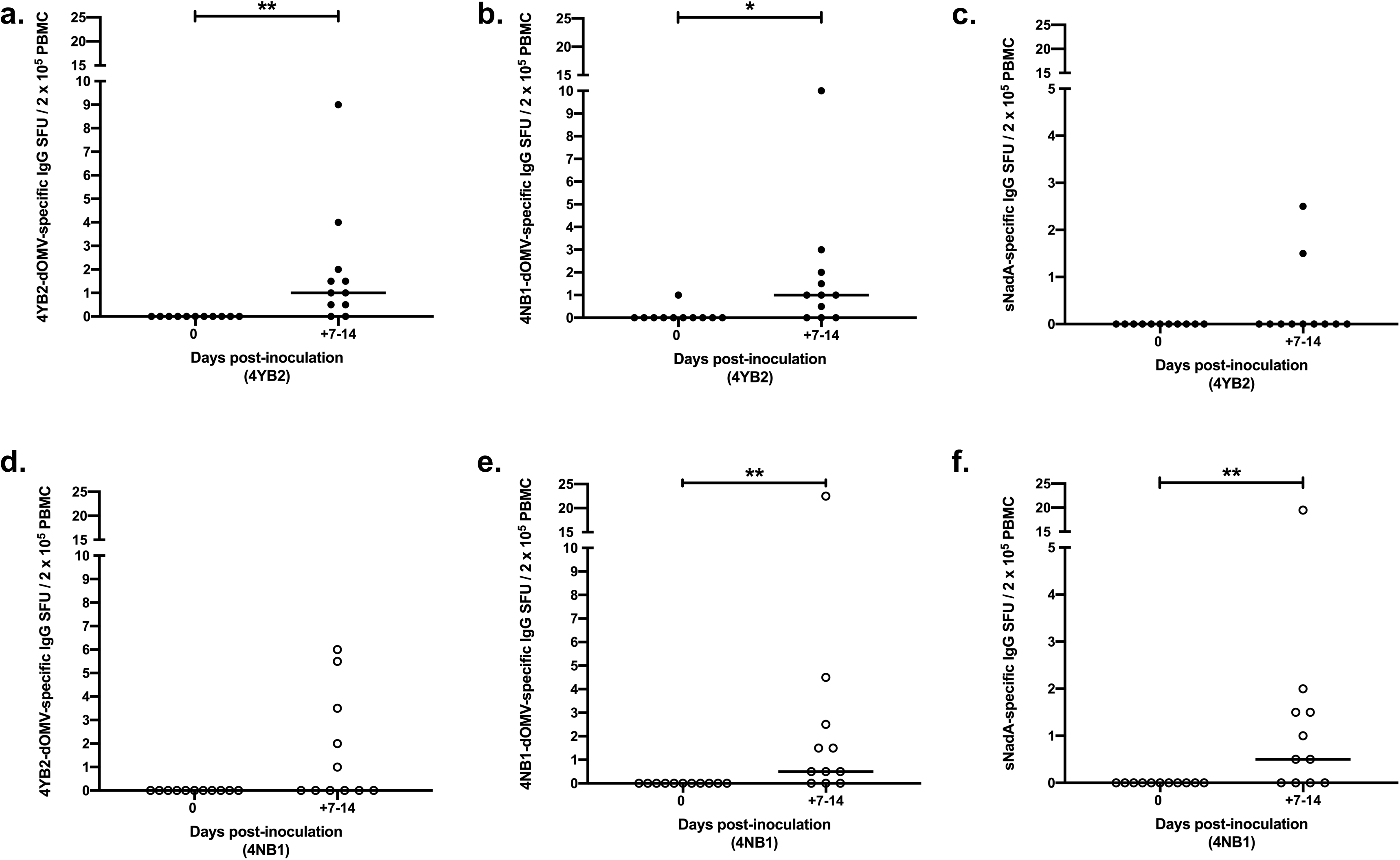
Colonization with NadA-expressing genetically modified *N. lactamica* results in the circulation of IgG-secreting plasma cells specific for NadA. PBMC isolated from whole blood at baseline and on Days 7 & 14 post-challenge with either control, WT-equivalent GM-Nlac (4YB2, •) or NadA-expressing GM-Nlac (4NB1, ○) were assessed by ELISpot for the presence of IgG-secreting cells specific for: 4YB2-dOMV (**a** and **d**), 4NB1-dOMV (**b** and **e**), sNadA (**c** and **f**) and tetanus toxoid (data not shown). IgG-secreting plasma cells were visualized as spot forming units (SFU) and adjusted for non-specific SFU by deducting the appropriate average number of SFU from KLH-coated membranes. Data are presented as the number of antigen-specific IgG SFU per 2 × 10^5^ PBMC. For each participant, the largest number of antigen-specific IgG SFU per 2 × 10^5^ PBMC (universally measured at either Day 7 or Day 14, i.e. the ‘peak’) is shown (+7-14). Bars indicate Median. \**p* ≤ 0.05, ** *p* ≤ 0.01 Wilcoxon matched-pairs signed rank test (n = 11).

### Colonization with NadA-expressing GM-Nlac elicits seroconversion against NadA

Colonization with either strain of GM-Nlac led to a significant and sustained increase in the interpolated reciprocal titer of anti-WT Nlac dOMV IgG (hereafter, anti-Nlac IgG) present in the sera of volunteers at Days 28, 56 and 90 compared to baseline (Figure 5). In 4YB2-colonized participants (Figure 5A), the median maximum fold change in anti-Nlac IgG was 3.05 (range: 1.5 – 16.9), whilst the same value was 2.8 (range: 1.4 – 12.8) in 4NB1-colonized participants (Figure 5B). In the three participants who were not colonized at any point by GM-Nlac, the median maximum fold change in interpolated reciprocal anti-Nlac dOMV IgG titer was 1.3 (range: 1.1 – 1.3) (data not shown). Anti-Nlac IgG calculated for sera taken from 4YB2-colonized participants was not significantly different to that from 4NB1-colonized participants at any point. Importantly, there was no correlation between baseline anti-Nlac IgG and maximum fold change (data not shown), which suggests this assay can measure increases in anti-Nlac IgG even in individuals with relatively high pre-existing titers of either Nlac-specific, or Nlac-cross reactive IgG. These data confirm that GM-Nlac colonized participants are generally responsive to the presence of the colonising bacteria and generate IgG against epitopes found on the surface of WT Nlac.

**Figure 5:**
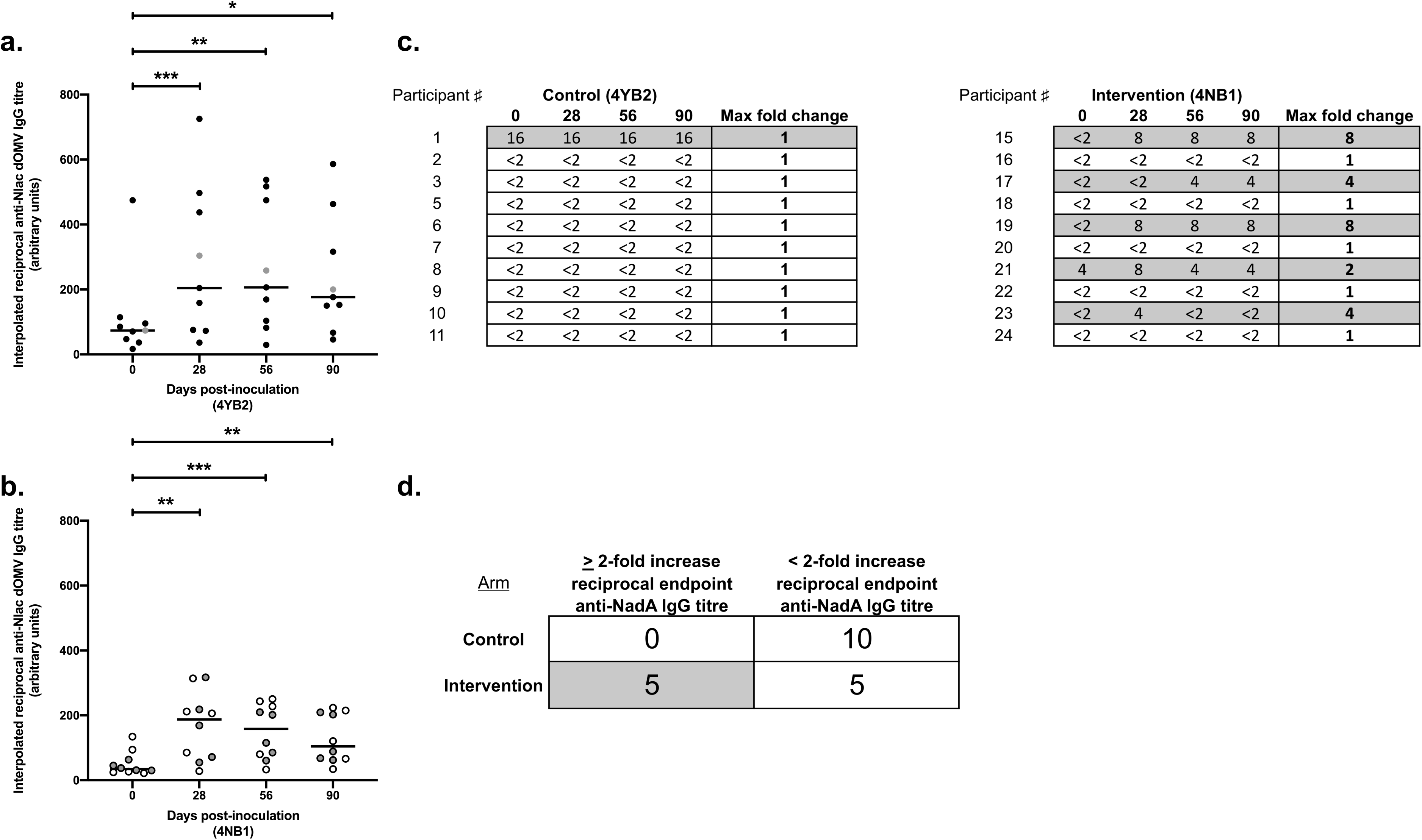
Colonization with genetically modified *N. lactamica* results in increased serological concentrations of IgG directed against *N. lactamica* surface epitopes, including NadA when present. Sera from volunteers colonised with (**a**) 4YB2 (Control, •) or (**b**) 4NB1 (Intervention, o) were assayed for IgG with specificity to epitopes present on WT Nlac dOMV (anti-Nlac dOMV IgG). Titers of anti-Nlac dOMV IgG in serum samples were interpolated with reference to serum NA9136. Bars denote Median. \**p* ≤ 0.05, \*\**p* ≤ 0.01, \*\*\**p* ≤ 0.001 Friedman’s 2-way Analysis of Variance by Ranks with Dunn’s multiple comparisons test vs. ‘0’ as control column. Only complete data sets from participants free from Nmen carriage at all time points were analysed (4YB2: n = 9, 4NB1: n = 10). Participants (numbered as in Figure 3B) in whose serum there was a detectable reciprocal endpoint titre of anti-NadA IgG (i.e. ≥ 2) at one or more time points are depicted/filled in grey (**a**, **b**, **c** and **d**). (**c**) Sera were assayed for anti-NadA IgG, using an endpoint ELISA. The reciprocal endpoint titer of each serum was considered to be the reciprocal titer of the least dilute serum sample tested that generated an OD_490nm_ ≥ 1.4. A reciprocal endpoint titer of anti-NadA IgG < 2 was considered to have a value of 1 for the purposes of calculating fold change, which was always in comparison to sera from Day 0. (**d**) Contingency table showing the numbers of participants from both arms of the study with either a ≥ 2-fold increase in the reciprocal endpoint anti-NadA IgG titer, or a < 2-fold increase (i.e. no detectable change) in the reciprocal endpoint anti-NadA IgG titer. Participants were considered to have a 2-fold increase if we measured a 2-fold increase in anti-NadA IgG in at least one post-baseline serum sample. *p* = 0.0325, Fisher’s exact test.

The titer of anti-sNadA IgG was also measured in each serum sample, using an endpoint enzyme-linked immunosorbent assay (ELISA) (endpoint = OD_490nm_ ≥ 1.4, see Supplementary Figure 11). Prior to inoculation, the majority of participants had undetectable levels of anti-sNadA IgG in their sera (reciprocal titer: ≤ 2), consistent with absence of colonization by Nmen at enrollment. It is likely that the majority of participants have previously been colonized by circulating strains of Nmen. However, because the *nadA* gene is only found in a subset of UK community meningococcal lineages^16^, there is only a small likelihood that participants would have generated immunological memory against NadA. Analysis of the maximum fold changes in anti-sNadA IgG shows that, whilst there is no increase in anti-sNadA IgG measurable in the sera of 4YB2-colonized participants; we measured at least a 2-fold rise in the reciprocal endpoint titer of anti-sNadA IgG at one or more time points in 5 of the 10 4NB1 - colonized participants (Figure 5C).

### Colonization with NadA-expressing GM-Nlac increases the circulating sNadA-specific memory B cell pool

IgG B cell memory to GM-Nlac and specifically to NadA was assessed by ELISpot assay^28^ (for representative outputs see Supplementary Figure 12). In all participants studied, no significant differences in the proportion of IgG memory B cells with specificity to influenza hemagglutinin (FluHA) (positive control antigen) were observed at any time point (Figures 6D & 6H) suggesting that the performance of the assay remained stable over time. These data suggest that any significant changes to the composition of the circulating memory B cell pool in response to Nlac-dOMV or sNadA were therefore specific to the experimental intervention.

**Figure 6:**
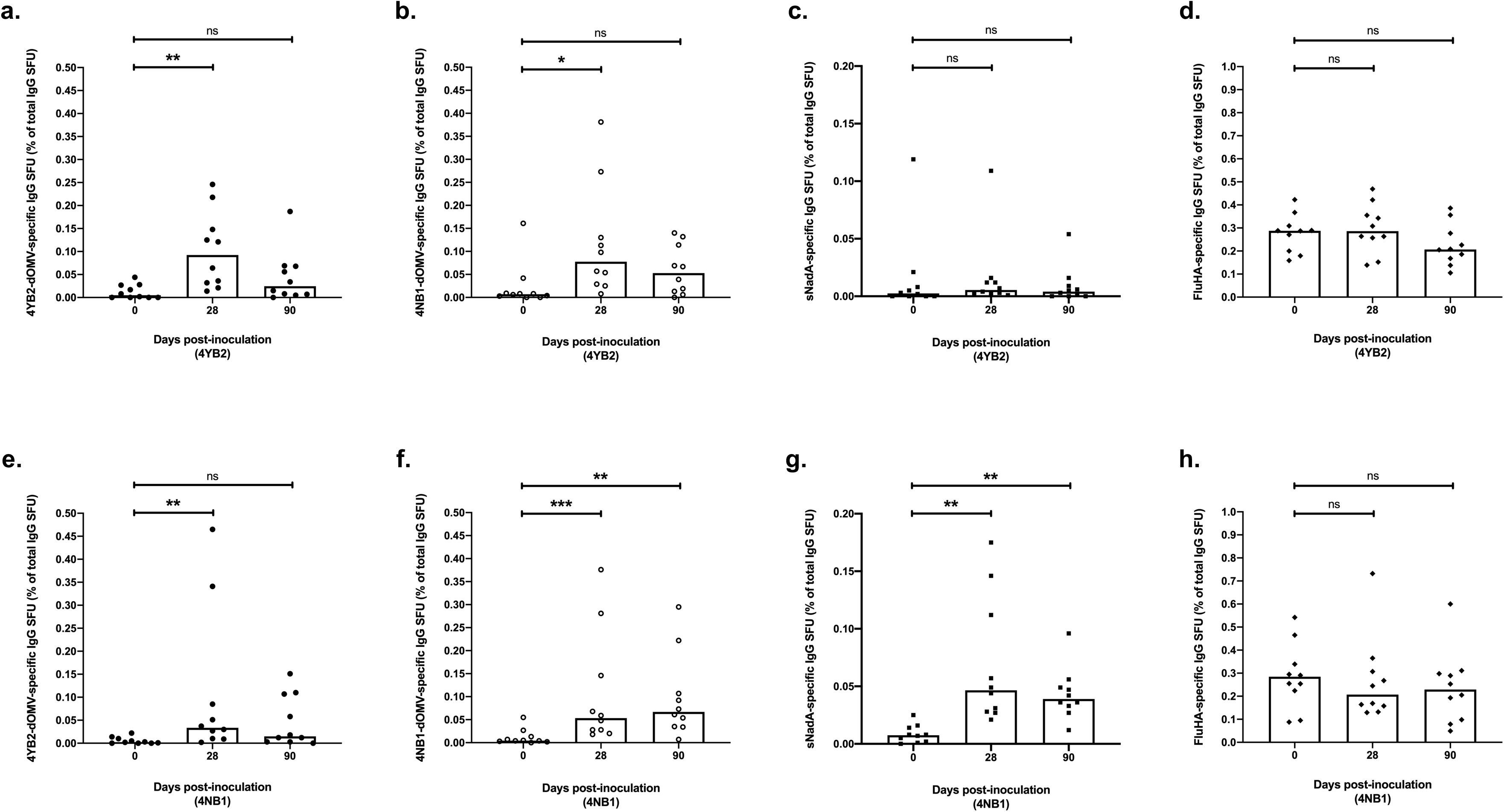
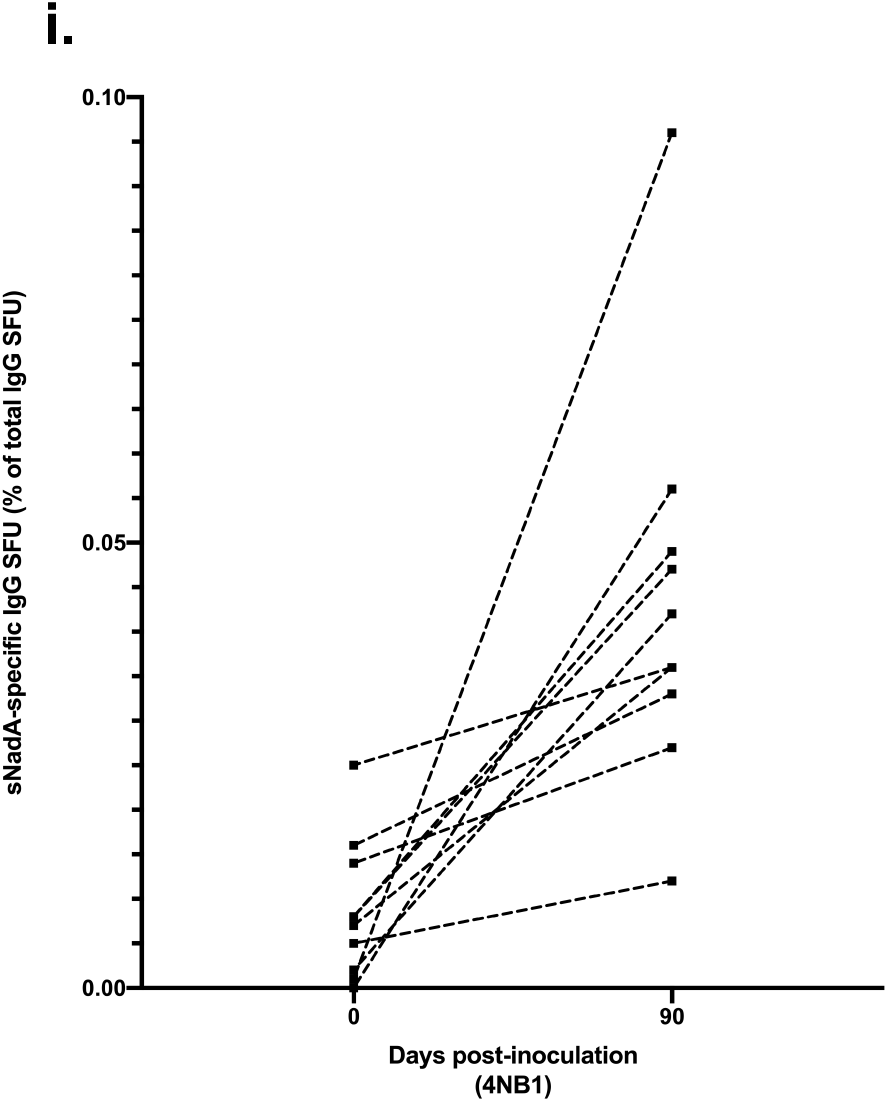
Colonization with genetically modified *N. lactamica* expressing NadA results in increased numbers of circulating, NadA-specific IgG memory B cells. PBMC isolated from whole blood at baseline and on Days 28 & 90 post-challenge with either control, WT-equivalent GM-Nlac (4YB2) or NadA-expressing GM-Nlac (4NB1) were polyclonally stimulated and assessed by ELISpot for the presence of IgG-secreting cells specific for: 4YB2-dOMV (•, **a** and **e**), 4NB1-dOMV (○, **b** and **f**), sNadA (■, **c**, **g** and **i**) and FluHA (control antigen) (♦, **d** and **h**). IgG-secreting cells were visualized as spot forming units (SFU) and averages were adjusted for non-specific SFU by deducting the appropriate average number of SFU from KLH-coated membranes. Antigen-specific IgG-secreting SFU are shown as a percentage of the total number of IgG-secreting SFU. Top bar of columns represents Median. ns: *p* > 0.05, \**p* ≤ 0.05, \*\**p* ≤ 0.01, \*\*\**p* ≤ 0.001, Friedman’s 2-way Analysis of Variance by Ranks with Dunn’s multiple comparisons test vs. ‘0’ as control column. (**i**) Percentages of NadA-specific SFU as proportions of the total number of IgG-secreting SFU at baseline and Day 90. Points derived from the same participant are linked by dashed lines.

In 4YB2-colonized participants, there was a significant but temporary increase in the percentage of the total number of IgG-secreting SFU (hereafter, % of memory SFU) with specificity to either 4YB2-dOMV or 4NB1-dOMV at Day 28, as compared to baseline. These increases were no longer evident at 90 days post-inoculation (Figures 6A & 6B, respectively). The same transient increase in the % of memory SFU specific for 4YB2-dOMV is evident in 4NB1-colonized participants (Figure 6E), but interestingly there was a significantly increased % of memory SFU specific for 4NB1-dOMV at Day 90 (Figure 6F).

In 4YB2-colonized participants there were no significant differences between the *%* of memory SFU with specificity to sNadA measured at any point (Figure 6C). In stark contrast, in 4NB1-colonized participants there were significant increases in the *%* of memory SFU with specificity to sNadA measured at both Day 28 and Day 90, as compared to baseline (Figure 6G). This suggests that the majority of SFU observed in 4NB1-dOMV-coated wells at Day 90 are binding to the NadA component of the dOMV, and that NadA-specific memory B cells continue to circulate in increased numbers (compared to baseline) in the bloodstream for at least 90 days post inoculation.

Importantly, in every participant that was colonized with strain 4NB1, there was a relative increase in the *%* of memory SFU with specificity to sNadA (Figure 6I).

## DISCUSSION

The remarkable finding in the current report is that, following a single, intranasal dose of a recombinant human commensal expressing a heterologous vaccine antigen, 100 % of colonized participants showed an increase in the proportion of circulating, antigen-specific memory B cells after 90 days. In contrast, participants colonized for the same length of time by an isogenic control bacterium experienced no substantial changes to the antigen-specific proportion of their memory B cell pool. In both cases, the inoculation was safe and colonization was sustained for 90 days, when participants received a carriage-clearing antibiotic.

Memory B cells do not have a primary antibody-secreting function, but promptly differentiate into plasma cells upon effective re-stimulation by cognate antigen^29^. This confers the potential to rapidly produce antibody following memory B-cell activation by a pathogen, such as one establishing pharyngeal colonization, preventing further progression to clinical disease. Commensurate with this, participants colonized by the NadA expressing strain of GM-Nlac had increased numbers of NadA-specific plasma cells and seroconverted to make detectable quantities of anti-NadA IgG. It is also important to note that increased numbers of memory B cells have been shown to be associated with resistance to nasopharyngeal bacterial colonization acquisition^30^. The data presented here suggests a vaccine-like potential for this technology, especially as a means to provoke anamnestic responses against established vaccine antigens.

This technology exploits the natural mechanism of protection against disease acquired by repeated or long-term colonization with pathobionts of the respiratory tract^1-3^. It does this by delivering antigen expressed by organisms replicating on the mucosal surface of the upper respiratory tract, effectively providing both antigen and adjuvant simultaneously. It is the only example of the use of a common human commensal for this purpose. The technology has many potential applications, as it is possible to introduce a wide variety of nucleic acid sequences into the bacterial chromosome, including codon-adjusted, exon-free eukaryotic genes (Supplementary Information). It is therefore theoretically compatible with technologies to express antigen or antigen fragments on the bacterial surface^31^.

This technology has a number of advantages over conventional bacterial vaccines. Firstly, the site of delivery is identical to the route of acquisition of upper airway pathogens. This is a property common to live attenuated influenza vaccine (LAIV), wherein poorly-replicating, attenuated virus, provided it is adequately matched^32^, is capable of inducing potent T-cell-mediated immunity against influenza virus when delivered intranasally, and consistently confers protective efficacy^32, 33^. Regarding bacterial vaccines, a Phase 1 trial of a live, genetically attenuated *Bordetella pertussis* intranasal vaccine has been reported. Transient, immunising asymptomatic colonization was demonstrated after inoculation with BPZE1, a *B.pertussis* strain in which dermonecrotic toxin and tracheal cytotoxin are both genetically deleted, and where the toxic activity but not the immunogenic property of pertussis toxin is genetically removed^34^. In mice, BPZE1 has been shown to induce an IL-17-dependent SIgA-mediated protection, likely due to the generation of tissue-resident memory T cells (T_RM_) that rapidly expand in the lungs and nasal cavity after infection and mediate rapid clearance of the pathogen^36^. The identification of T_RM_ cells and their protective capacity for tissue-specific protection from infection is a powerful new tool for vaccine design^37-39^ that is harnessed by recombinant commensal infection. In addition, the technology reported herein, in contrast to live attenuated vaccines, confers safety not by attenuation, but by use of a common commensal of infants as the bacterial vector.

Secondly, the immunity conferred by sustained carriage of the bacterial vector is likely to be protracted. Although the longevity of the NadA-specific IgG memory B cells in the circulation beyond 90 days has not been established, this could be investigated in future work. The currently deployed 4CMenB vaccine, which contains NadA, generates relatively short-lived immunity even after boosting, and its effects on Nmen colonization and therefore herd immunity may be limited^40^. Although the scope of the technology need not be limited to meningococcal disease; in the specific context of novel meningococcal vaccines, this approach could be further developed to induce immunological responses towards heterologous meningococcal antigens, in order to provide protection against invasive meningococcal disease, with additional herd immunity effects provided as a consequence of the aforementioned competition for niche occupation between Nlac and Nmen^6^.

In conclusion, we describe the safety and potential utility of a genetically transformed nasopharyngeal commensal to induce long lasting humoral immunity to heterologous antigen in humans.

## METHODS

### Bacterial isolates and routine culture methods

All stocks of Nlac, Nmen, and their mutant derivatives (Supplementary Table 1) were maintained as frozen glycerol stocks at −80°C. Upon recovery from cryostorage Nlac was cultured on tryptone soya agar (TSA) supplemented with X-Gal (40 μg ml^-1^) where appropriate. Nmen was cultured on Columbia agar supplemented with horse blood (5 % v/v) (CBA), supplemented with kanamycin (50 μg ml^-1^) where appropriate. Routine liquid culture of Nlac was performed in TSB, at 37°C, 5 % CO_2_.

Nmen serogroup B strain MC58 and its mutant derivatives (MC58*∆nadA* & MC58*∆siaD*) were cultured in Mueller-Hinton broth (MHB), supplemented with kanamycin (50 μg ml^-1^). Nmen serogroup Y strain N54.1 and its mutant derivative (N54.1*∆nadA*) were cultured in MHB supplemented with 5 mM 4-hydroxyphenylacetic acid (4HPA) (Sigma-Aldrich, cat no. H50004-5G). Nmen NadA-overexpressing strain 5/99 was cultured in modified Frantz medium^41^ For determination of clinically relevant antibiotic MICs, Nlac was cultured as a lawn on Mueller-Hinton agar supplemented with horse blood (5 % v/v) in the presence of the appropriate e-test strips: ciprofloxacin and ceftriaxone (Biomerieux). Note that as no interpretative criteria exist for determining antibiotic susceptibility of Nlac (WT or GM), two reference criteria were used. The first was the European Committee on Antimicrobial Susceptibility Testing (EUCAST) breakpoints for *N. meningitidis* and the effective treatment of meningococcal disease and the second was the antibiotic susceptibility profile of 286 isolates of WT *N. lactamica*^26^

### Detection and identification of *Neisseria* species in clinical samples

*Neisseria* species were isolated from clinical samples using GC selective agar plates, supplemented with 10 % (v/v) lysed horse blood, VCAT selective supplement (Oxoid, cat no. SR0104) (1 % v/v), Vitox supplement (Oxoid, cat no. SR0090) (2 % v/v), glucose (0.4 % w/v), Amphotericin B (10 μg ml^-1^) and X-gal (40 μg ml^-1^) (GC+Xgal) (Southern Group Laboratories). Pre-clinical tests demonstrated that only Nlac grew as blue coloured colonies, while all other *Neisseria* species tested grew as white colonies (data not shown). For each participant at each time point, up to 10 colonies (preferentially 5 from nasal wash, 5 from throat swab) of blue, putative GM-Nlac were subcultured onto fresh GC+X-gal plates for the preparation of stocks and lysates for diagnostic PCR. White-coloured, putative Nmen colonies were tested for oxidase activity using oxidase detection strips (Oxoid, cat no. MB0266). All oxidase positive, Gram-negative diplococci were subcultured onto fresh Columbia agar plates supplemented with chocolated horse blood (5 % v/v) (CHOC), incubated overnight at 37°C, 5 % CO_2_ and then used to produce a cell suspension suitable for identification using API NH kits (Biomerieux, cat no. 10400), performed according to the manufacturer’s instructions. Bacterial stocks were created of all carriage Nmen isolates identified during the study.

### Culture methods for preparation of human challenge inocula

Inocula preparation was carried out to Good Manufacturing Practice (GMP)-like standards in a Containment Level 2 (CL2) laboratory at the University of Southampton. Access to the CL2 laboratory was restricted to all but essential, fully trained personnel. Inocula of GM-Nlac strains, 4NB1 and 4YB2 were prepared separately, with all stages of the production process for one strain (4YB2) being completed before commencing production of the second (4NB1). Prior to and between production runs, all laboratory equipment was decontaminated or sterilized. To prepare the inocula, seed cultures of the appropriate GM-Nlac strain were grown overnight at 37°C, 5 % CO_2_ on TSA plates supplemented with X-gal. Blue-colored GM-Nlac colonies were subcultured in TSB (Vegitone) at 37°C, 5 % CO_2_ to an OD_600nm_ of 1.5 arbitrary units. Subcultures were assessed for NadA expression by flow cytometry and then used to prepare the ‘Master Stock’. Briefly, bacteria were pelleted by centrifugation (10 min @ 5,000 *g)*, washed by resuspension in PBS, and then resuspended in Frantz medium:glycerol (70 % : 30 % v/v). Aliquots were stored in numbered ‘Master Stock’ cryovials at −80°C. Master stock viability and purity was determined by culture on TSA, CBA and CHOC plates overnight at 37°C, 5 % CO_2_. Subsequently, contaminant-free ‘Master Stock’ vials were diluted into ice-cold Frantz medium:glycerol (70% : 30% v/v) to a final concentration of 5 × 10^6^ CFU ml^-1^. One-millilitre aliquots were then stored in numbered ‘Inoculum’ cryovials at −80°C. By culturing on ‘contaminant-free’ CBA plates, the average viability of the ‘Inoculum’ vials was calculated as 4.8 × 10^6^ CFU ml^-1^ for strain 4YB2 (n = 7) and 4.8 × 10^6^ CFU ml^-1^ (n = 8) for strain 4NB1.

### Preparation of inoculum for intranasal administration

The preparation of inocula for administration into human participants was performed in a class 2 microbiological safety cabinet inside a CL2 laboratory in the CRF, Southampton General Hospital. Briefly, a 1 ml ‘Inoculum’ vial containing the appropriate GM-Nlac strain was thawed at RT, vortexed, and 500 μl was diluted into 4.5 ml sterile PBS pH 7.4 (Severn Biotech). Intranasal inoculation of participants occurred within 30 min of defrosting the vial. The purity and viability of the inoculation dose was determined by culture on CHOC agar.

### Preparation of gDNA

Bacterial strains were cultured to mid-log phase in an appropriate liquid culture medium, supplemented where appropriate with the relevant antibiotics. Bacterial gDNA was harvested using a GeneJET Genomic DNA Purification kit (Thermofisher, cat no. K0722) according to Protocol D (Gram-negative bacteria) in the manufacturer’s instructions. The concentration of gDNA was measured using a Nanodrop spectrophotometer (Shimadzu).

### Transformation of Neisseria species

WT Nlac strains Y92-1009, 4NB1 and WT Nmen strain MC58 were all grown in liquid culture to mid-log phase. Bacteria were pelleted, washed in PBS and resuspended in TSB supplemented with 10 mM MgSO_4_ at an OD_600nm_ = 0.3. One milliliter aliquots of each bacterial suspension were incubated at 37°C, 5 % CO_2_ for 3 h with 1 × 10^4^ pmoles of gDNA harvested from one of the following Nlac or Nmen mutant strains: (i) Nlac strain Y92-1009*∆nlaIII:aphA3*, (ii) Nmen strain MC58*∆AnadA:aphA3*, (iii) Nmen strain MC58*∆siaD:aphA3*. Aliquots of each suspension were spread on CBA plates supplemented with kanamycin (50 μg ml^-1^). The total viability (in CFU ml^-1^) of each suspension was determined through serial dilution and plating on CBA. Transformation efficiency was calculated by dividing the number of colonies of transformed (i.e. kan^R^) bacteria (in CFU ml^-1^) by the total number of bacteria in the suspension (in CFU ml^-1^), and normalizing to 1 pmol gDNA.

### Diagnostic Polymerase Chain Reaction (PCR)

Bacterial isolates from clinical samples were identified as GM-Nlac using multiplex PCR. Briefly, lysates from putative GM-Nlac colonies were used as template DNA to amplify a GM signature sequence (Band3d, 284 bp) and/or the 16S rRNA gene (Band1, 469 bp). Genomic DNA from GM-Nlac strain 4NB1 (50 ng ml^-1^) was used as a Band3d positive control, gDNA from WT Nmen strain H44/76 (50 ng ml^-1^) was used as a Band1 positive control, and RNAse-free H_2_O was used as a negative control. Each PCR reaction volume (20 μl) comprised one of control gDNA, test lysate or RNAse-free H_2_O (1 μl), plus Platinum II Taq Hot-Start DNA polymerase Master mix (2x) (Thermofisher, cat no. 14966025) (10 μl), Band1FOR (10 μM) (0.8 μl), Band1 REV (10 μM) (0.8 μl), Band3dFOR (5 μM) (0.4 μl), Band3dREV (5 μM) (0.4 μl) and RNAse-free H_2_O (6.6 μl). PCR amplification used the following, 2-step thermal cycling parameters: [1 × 2 min @ 95°C] followed by [35 × (5 sec @ 98°C, 15 sec @ 60°C and 15 sec @ 68°C)]. Amplicons were analysed by 2% agarose gel electrophoresis and visualized under UV transillumination using a Gel Doc XR+ (Bio Rad). Preliminary work using both purified gDNA and bacterial lysates showed that amplification of Band3d often resulted in no little to no amplification of Band 1 in a multiplex reaction, but that in the absence of the GM signature sequence, Band1 was the only amplification product (data not shown). Therefore, GM-Nlac was positively identified if either a single amplicon of 284bp, or two amplicons of 284 bp and 469 bp were present. A single amplicon of 469 bp was determined to be something other than GM-Nlac. PCR yielding zero amplicons was considered to have failed, and was repeated until definitively identified as being, or not being, GM-Nlac.

### Flow cytometry

Surface expression of NadA on Neisseria spp. was assessed by flow cytometry of fixed cells. Briefly, bacteria (2 × 10^7^ CFU) were labeled with either anti-NadA mAb 6e3^42^ or anti-NadA pAb (GSK) in wash buffer (PBS + 5 % FCS) for 30 min at 4°C. Cells were washed and stained with AlexaFluor488-labeled goat anti-mouse IgG antibody in wash buffer for 30 min at 4°C, and then fixed in formalin for 15 min. Bacteria were gated based upon forward and side scatter profiles and a total of 10,000 events were acquired on FacsCalibur (BD Biosciences). Relative NadA expression levels were measured as the Mean Fluorescence Intensity (MFI) of Alexafluor488, recorded in the FL-1 channel for the gated events.

### Adherence assay

HEP-2 cells (2 × 10^5^ cells well^-1^) were seeded into 24-well plates and cultured for 2 days in Dulbecco’s Modified Eagle’s medium (DMEM) supplemented with 10 % Foetal Calf Serum (FCS) at 37°C, 5 % CO_2_. HEP-2 cells were then washed twice in sterile PBS, then infected at MOI = 100 with the relevant bacterial strain. Plates were incubated at 37°C, 5 % CO_2_ and at 2 h, 4 h and 6 h, supernatants were aspirated and wells washed five times with PBS. Two percent saponin in PBS (250 μl well^-1^) was added and plates were incubated for 15 min at 37°C, 5 % CO_2_. Wells were supplemented with PBS to a final volume of 1 ml and cells were mechanically agitated through pipetting. The number of viable bacteria was determined by plating serially diluted saponised cell lysate onto CBA plates. The viability of each lysate was normalised to the number of HEP-2 cells per well, which was estimated from the cell count of duplicate wells.

### Isolation of Outer Membrane Vesicles

OMV were deoxycholate-extracted from bacterial cell pellets grown in modified Caitlin (MC7) medium^41^. Briefly, *Neisseria* were cultured overnight at 37°C, 5 % CO_2_ to OD_600nm_ > 2.0 and pelleted by centrifugation at 4500 *g* for 1 h. Cell pellets were resuspended using a glass homogeniser into 0.1 M Tris-HCl (pH 8.6) with 10 mM EDTA and 0.5 % (w/v) deoxycholic acid sodium salt. These suspensions were centrifuged at 20,000 *g* for 30 min at 4°C, and the supernatant was retained. The above resuspension/homogenization procedure was repeated once to maximize yield. OMV were harvested from the combined supernatants by ultracentrifugation at 100,000 *g* for 2 h at 4°C. Pelleted OMV were gently resuspended into 50 mM Tris-HCl (pH 8.6) with 2 mM EDTA, 20 % (w/v) sucrose and 1.2 % deoxycholic acid sodium salt. OMV were again pelleted by ultracentrifugation at 100,000 *g* for 2 h at 4°C, before resuspension into 50 mM Tris-HCl with 3 % sucrose, which formed the final formulation. The protein content of the OMV formulation was measured by the DC Protein assay (Biorad), with reference to a standard curve comprised of known bovine serum albumin (BSA) concentrations.

### Murine Immunisations with dOMV

Groups of ten 6-8-week-old BALB/c mice were immunized by subcutaneous injection with three doses of 10 μg Nlac dOMVs containing 0.33% Alhydrogel^43^ at day 0, 21 and 28. Sera were collected on day 35.

### Murine intraperitoneal (i/p) challenge model

WT Nlac strain Y92-1009 and the GM-Nlac strains 4YB2 and 4NB1 were cultured in TSB medium to mid-log phase, washed in PBS, and resuspended at different concentrations into 50 μl PBS supplemented with 10 mg of human holo-Transferrin (Sigma-Aldrich, cat no. T4132). Six groups of NIH/OLA mice (6-8 weeks), comprised of 5 male and 5 female mice per group, were challenged with ‘high’ or ‘low’ i/p doses of the Nlac/GM-Nlac strains. Viability of each dose was determined by colony count on CBA plate cultures. Twenty-four hours after challenge, the mice were given an additional i/p dose of 10 mg human holo-Transferrin. The health of the mice was assessed based on their outward appearance and behaviour and was recorded every 4 h in the first 2 days and then every 6-8 h up to 5 days post-challenge. Behaviourally ‘normal’ mice were considered ‘healthy’ (recorded as ‘H’), whilst those experiencing mild discomfort presented as ‘arched’ (recorded as ‘A’) and/or ‘ruffled’ (recorded as ‘R’).

### Serum bactericidal assays

SBA assays were performed against Nmen strain 5/99, resuspended in bactericidal buffer (BB; Hanks buffered saline solution (Invitrogen) and 1% BSA) at 6 × 10^4^ CFU ml^-1^. Twofold dilutions of heat-inactivated sera were prepared in microtiter plates (20 μl), to which Nmen (10 μl) was added followed by IgG-depleted pooled human plasma as an exogenous human complement source (10 μl) (i.e. 25% final concentration). Plates were incubated at 37°C for 1 h with shaking at 900 rpm. Each sample and control well was then plated onto CBA using the tilt method and incubated overnight at 37°C, 5% CO_2_. The following day colonies were counted and % survival was determined in comparison to colony numbers in the t = zero control. Dilutions of sera were deemed bactericidal if bacterial survival rates were ≤ 50 %. SBA reciprocal titres were determined as the reciprocal of the most diluted serum sample that was bactericidal.

### Recombinant expression of the soluble domain of NadA

Recombinant expression of sNadA was carried out commercially by BioServUK (Sheffield, UK), broadly consistent with the methodology described by Cecchini and colleagues^44^. Briefly, the coding sequence for *nadA* allele 1, minus the sequence coding for the C-terminal YadA domain *(nadA∆285-364)*, was cloned from pJL0017 into pRSETα to include a C-terminal 6His-tag. Protein secreted into growth medium by the transformed *E. coli* strain BL21 (DE3) was purified using HiFliQ column chromatography and buffer exchanged into PBS (pH7.5). Analysis of purified sNadA by SDS-PAGE showed a single band by silver staining under reducing and denaturing conditions, and three bands, consistent with homotrimer formation, under denaturing but non-reducing conditions. Purified sNadA was cross-reactive with anti-NadA mAb 6e3 (GSK) in western blotting analysis. Purified sNadA was stored at −80°C and diluted in PBS to make working stocks (1 mg ml^-1^), which were stored at −20°C.

### ELISA for measurement of human anti-Nlac IgG

Nunc Maxisorb 96-well plates were coated overnight at 5°C with WT Nlac Y92-1009 dOMVs (10 μg ml^-1^) in carbonate coating buffer (pH 9.6). Coated plates were washed five times with PBS containing 1 % Tween 20 (PBST) (Skatron SkanWasher 400) and blocked with PBST containing 5 % (v/v) FCS for 90 min at RT. Plates were washed five times with PBST and then loaded in duplicate with serially diluted test samples and reference serum (NA9136). Dilutions were prepared in Sterilin Serowell (low binding) plates with a starting dilution of 1/25 followed by three-fold serial dilution across the plate. Loaded plates were incubated with shaking at RT for 90 min. Plates were then washed five times with PBST prior to incubation at RT for 90 min with goat anti-human IgG Fcγ-fragment-specific alkaline phosphatase conjugate (Jackson Immunoresearch Laboratories cat no. 109-055-098) diluted 1/1750 in PBST + 5 % (v/v) FCS. This was followed by a wash step as above, and addition of AP yellow (p-nitrophenyl phosphate) substrate (BioFX Laboratories, Cat. No. PNPP-0100) for 55 min at RT. The reaction was stopped with 3M NaOH. Optical density was read using a VersaMax plate reader at 405nm with a reference wavelength of 690nm. SoftMax® PRO Enterprise software (Molecular Devices) was used to fit an un-weighted five parameter logistic (5PL) log curve to titrations of the reference serum for each plate. Test sample titres were calculated by interpolation from the reference serum dose response curve (interpolated titre multiplied by dilution factor). A mean of the acceptable values from all dilutions was then taken as the final value for each test serum.

### ELISA for measurement of human anti-sNadA IgG endpoint titers

Corning Costar EIA/RIA 96 well plates were coated for up to 1 week at 4°C with sNadA (10 μg ml^-1^) in carbonate coating buffer (pH 9.6). Coated plates were washed three times with 250 μl sterile PBS and blocked with 5 *%* FCS-PBS at 37°C for 2 h. Plates were washed four times with 250 μl PBS and then loaded (in duplicate) with twofold dilutions of test sera (50 μl). Control serum (NA9136) was diluted 1/16 in 5 *%* FCS-PBS, as this dilution was shown to generate a positive signal under the conditions used in this ELISA (Supplementary Figures).

Loaded plates were incubated at 37°C, 5 % CO_2_ for 1 h after which they were washed five times with PBS supplemented with 0.05 *%* Tween 20 (hereafter, PBS-T). Biotinylated, Rat-derived anti-human IgG mAb (M1310G05) (Biolegend, cat no. 410718) was diluted 1/1000 in 5 *%* FCS-PBS and 50 μl was added to each well. Plates were incubated at 37°C, 5 *%* CO_2_ for 1 h, after which they were washed five times with PBS-T. A streptavidin-horse radish peroxidase (HRP) conjugate (Biolegend, cat no. 405210) was also diluted 1/1000 in 5 *%* FCS-PBS and added to each well (50 μl). Plates were incubated at 37°C, 5 % CO_2_ for 1 h, after which plates were washed five times with PBS-T and a single wash with PBS. Chromogenic substrate, o-phenylenediamine dihydrochloride (OPD) (Thermofisher, cat no. 34006), was prepared by dissolving 1 OPD tablet into 9 ml of dH_2_O supplemented with 1 ml stable peroxide substrate buffer (Thermofisher, cat no. 34062). The plate was incubated with OPD substrate (100 μl well^-1^) at 32°C for exactly 20 min. Signal development was stopped with 2 N H_2_SO_4_ (50 μl well^-1^), and the color-change quantified on a Versamax plate reader (Molecular Devices) measuring at optical density 490nm (OD_490nm_). The reciprocal endpoint titre of each test serum was considered to be the reciprocal titre of the least diluted serum sample tested that generated an OD_490nm_ in excess of 1.4 arbitrary units in this assay.

### Isolation of peripheral blood mononuclear cells

PBMC were isolated from whole blood of CHIME participants by density gradient centrifugation and used immediately in the plasma cell ELISpot assay. The remainder were stored in LN_2_ for later use in the memory B cell ELISpot assay.

### Polyclonal stimulation of peripheral blood mononuclear cells

PBMC were cultured and polyclonally stimulated as outlined previously^45^. Briefly, PBMC isolated from the whole blood of participants taken at Day 0, Day 28 and Day 90 visits were cultured separately in 96-well tissue culture plates (ThermoFisher) at 2 × 10^5^ cells per well in 200 μl AIM/V + albumax medium (Gibco, Invitrogen) supplemented with 10 % FCS and 50 pM β-mercaptoethanol (hereafter, AIM/V+). Cultures were supplemented with 3 μg ml^-1^ human phosphorothioate-modified oligodeoxynucleotide containing CpG motifs (ODN2006: 5’-TCG TCG TTT TGT CGT TTT GTC GTT-3’) (InvivoGen), 10 ng ml^-1^ IL-2 (R&D Systems), and 10 ng ml^-1^ IL-10 (Pharmingen, BD), and incubated for 5 days at 37°C, 5 % CO_2_.

### ELISPOT Assay

Ninety-six-well ELISpot plates (Multiscreen HTS plate, Merck Millipore, cat no. MSIPS4510) were activated with 70 % (v/v) ethanol, washed three times with PBS and coated with 100 μl PBS containing one of the following antigens: keyhole limpet hemocyanin (KLH) (Sigma-Aldrich) (10 μg ml^-1^), tetanus toxoid (TT) (2.5 level of flocculation (LOF) units /ml), influenza hemagglutinin (FluHA) (influenza antigen reagent 09/174, H1N1, NIBSC, UK) (0.5 μg ml^-1^), dOMV derived from GM-Nlac strain 4YB2 (4YB2-dOMV) (10 μg ml^-1^), dOMV derived from GM-Nlac strain 4NB1 (4NB1-dOMV) (10 μg ml^-1^), sNadA (10 μg ml^-1^) or rat-anti-human IgG mAb (clone M1310G05, IgG2a, k, Biolegend) (anti-IgG mAb) (10 μg ml^-1^). All plates were stored at 4°C overnight or maximally up to a week. Prior to use, plates were washed four times with PBS and blocked by incubation at 37°C for 2 h with AIM-V medium supplemented with 10 % FCS. Blocked plates were washed four times with PBS, before adding PBMC to each well and incubating for 18 h at 37°C, 5 % CO_2_. For the plasma cell ELISPOT, PBMC were freshly isolated and plated in duplicate at 2 × 10^5^ cells well^-1^ in AIM-V medium + 10 % FCS. For the memory B cell assay, polyclonally-stimulated PBMC (see above) were harvested and washed in AIM/V+ media by centrifugation at 300 g. Cells were resuspended in AIM/V+ medium at a concentration of 5 × 10^6^ cells ml^-1^, serially diluted, and seeded into ELISpot plate wells (200 μl volume) in duplicate or triplicate at concentrations of 5 × 10^6^ cells ml^-1^, 2 × 10^6^ cells ml^-1^, 1 × 10^6^ cells ml^-1^ and 2.5 × 10^5^ cells ml^-1^.

Following incubation, plates were washed four times with PBS-T and cells were lysed by washing three times with dH_2_O, followed by a 5 min incubation at RT in dH_2_O. Plates were washed as before and incubated for 1 h at 37°C, 5 % CO_2_, with alkaline phosphatase-conjugated anti-human IgG pAb (Sigma-Aldrich), prepared in PBS supplemented with 0.01 % Tween 20 and 1 % goat serum (Binding Buffer). Unbound pAb was removed by washing with PBS-T, rinsing the backs of the PVDF membranes gently with tap water and a final three washes with PBS. Plates were then incubated at RT with 50 μl well^-1^ BCIP substrate solution (prepared from SigmaFAST BCIP tablets in dH_2_O, Sigma cat no. B5655) for 13 min, before washing with tap water. ELISpot plates were dried and imaged using an AID ELISpot reader. IgG spot-forming units (SFU) were counted using the AID ELISpot software package, version 3.5. For the memory B cell assay, antigen-specific IgG SFU frequencies were derived for NadA, 4NB1-dOMV, and 4YB2-dOMV-coated wells from the lowest input cell concentration where a mean of 1-20 IgG SFU were counted following subtraction of KLH background. These antigen-specific SFU were expressed as a percentage of total IgG SFUs, as described previously^46^.

### Statistical Plan

Standard deviations (SDs) associated with serological responses to WT Nlac derived from our first Nlac CHIME were utilised to inform the sample size calculation^5^. This gave SDs on a log-10 scale of 0.26 for serum IgG. Using the SD of 0.26, it was calculated that a 4-fold rise in Nlac-specific IgG titer would be confirmed with 10 carriers of GM-Nlac expressing NadA with 90% power using analysis of variance.

All statistical analyses were performed using GraphPad Prism software (version 8.0). Parametrically- and non-parametrically-distributed continuous variables were summarised with mean ± standard deviation (SD) or median, respectively, and analysed using an appropriate parametric or non-parametric test, as indicated in each figure legend. The proportion of participants with ≥ 2-fold rise in anti-NadA IgG titer in control vs. intervention groups was compared using Fisher’s exact test. All *p* values were two-tailed and *p* values ≤ 0.05 were considered statistically significant.

## Data Availability

The complete reference genome of parent strain Y92-1009 is available as published work and on GenBank (accession: CP019894.1). Other data supporting the findings of the study are available in this article and its Supplementary Information file, or from the corresponding author upon request.

## ACKNOWLEDGEMENTS

We thank all of our study participants and the staff at the NIHR Clinical Research Facility at Southampton General Hospital for delivery of the clinical component of this study, notably Nursing Lead Sarah Horswill, Human Challenge Coordinator Sara Hughes and the CRF laboratory support staff. We thank Caroline Barker and the members of the Southampton Public and Patient Involvement group for their assistance in shaping the national press release that preceded our application to DEFRA. We thank Martin Cannell and Ivy Wellman from DEFRA and Richard Lockey from University of Southampton for guiding us through the Deliberate Release application process. We also thank Mariagrazia Pizza and colleagues at GSK for antibody reagents, and Chris Bayliss for kindly providing meningococcal strain N54.1.

This work was funded by the UK Medical Research Council (MR/N013204/1: ‘A genetically modified nasopharyngeal commensal as a platform for bacterial therapy’, and MR/N026993/1: ‘Pathfinder: Experimental Human Challenge with Genetically Modified Commensals to Investigate Respiratory Tract Mucosal Immunity and Colonization’) with additional funding from the MRC Confidence in Concept Award and the Wessex Institute for Vaccines and Infectious Disease. Robert Read is an NIHR Senior Investigator (NF-SI-0617-10010), Adam Dale is supported by the Wellcome Trust Research Training Fellowship (203581/Z/16/Z) and the work is supported by the National Institute for Health Research Southampton Biomedical Research Centre (IS-BRC-1215-20004).

## Contributions

J.R.L., D.G., A.D., M.C.M., S.F., A.G. and R.C.R. designed the study. D.G., H. D.G., S.F. and R.C.R. wrote the clinical protocol. J.R.L. developed the Nlac transformation technology. J.R.L., Z.P., G.B., K.B., D.W.C., A.P. and H.H. performed the pre-clinical laboratory experiments. J.R.L., C.W. and E.R. manufactured the challenge inocula. D.G., J.R.L., C.W., E.R., M.I. and A.R.H conducted or supported the clinical study. H.H., L.A., J.R.L and A.G. performed serological analyses and analysed the serological data. A.D., E.R., A.R.H. and J.R.L. performed cellular experiments and analysed the cellular data. J.R.L, A.D., A.R.H., D.G. and R.C.R. wrote the paper.

## ETHICS DECLARATIONS

### Ethical approvals and trial registration

This study has been approved by the Department for Environment, Food and Rural Affairs (reference: 17/R50/01) and South Central Oxford A Research Ethics Committee (reference: 18/SC/0133).

Informed consent was obtained from all participants prior to their enrolment in the controlled human infection model experiment.

Trial registration number: <u>NCT03630250</u>

Animal experiments were approved by the Public Health England, Porton Down Ethical Review Committee and authorised under an appropriate UK Home Office project license.

## REFERENCES

1. Goldblatt, D. et al. Antibody responses to nasopharyngeal carriage of *Streptococcus pneumoniae* in adults: a longitudinal household study. J Infect Dis 192, 387–393 (2005).

2. Hall, D.B., Lum, M.K., Knutson, L.R., Heyward, W.L. & Ward, J.I. Pharyngeal carriage and acquisition of anticapsular antibody to *Haemophilus influenzae* type b in a high-risk population in southwestern Alaska. Am J Epidemiol 126, 1190–1197 (1987).

3. Jones, G.R. et al. Dynamics of carriage of *Neisseria meningitidis* in a group of military recruits: subtype stability and specificity of the immune response following colonization. J Infect Dis 178, 451–459 (1998).

4. Gold, R., Goldschneider, I., Lepow, M.L., Draper, T.F. & Randolph, M. Carriage of *Neisseria meningitidis* and *Neisseria lactamica* in infants and children. J Infect Dis 137, 112–121 (1978).

5. Evans, C.M. et al. Nasopharyngeal colonization by *Neisseria lactamica* and induction of protective immunity against *Neisseria meningitidis*. Clin Infect Dis 52, 70–77 (2011).

6. Deasy, A.M. et al. Nasal Inoculation of the Commensal *Neisseria lactamica* Inhibits Carriage of Neisseria meningitidis by Young Adults: A Controlled Human Infection Study. Clin Infect Dis 60, 1512–1520 (2015).

7. Pandey, A. et al. Microevolution of *Neisseria lactamica* during nasopharyngeal colonisation induced by controlled human infection. Nat Commun 9, 4753 (2018).

8. Sardinas, G., Reddin, K., Pajon, R. & Gorringe, A. Outer membrane vesicles of *Neisseria lactamica* as a potential mucosal adjuvant. Vaccine 24, 206–214 (2006).

9. Chi, H. et al. Engineering and modification of microbial chassis for systems and synthetic biology. Synth Syst Biotechnol 4, 25–33 (2019).

10. Michon, C., Langella, P., Eijsink, V.G., Mathiesen, G. & Chatel, J.M. Display of recombinant proteins at the surface of lactic acid bacteria: strategies and applications. Microb Cell Fact 15, 70 (2016).

11. Zhou, Z. et al. Engineering probiotics as living diagnostics and therapeutics for improving human health. Microb Cell Fact 19, 56 (2020).

12. O’Dwyer C, A. et al. Expression of heterologous antigens in commensal *Neisseria* spp.: preservation of conformational epitopes with vaccine potential. Infect Immun 72, 6511–6518 (2004).

13. Pandey, A.K. et al. Correction to: *Neisseria lactamica* Y92–1009 complete genome sequence. Stand Genomic Sci 13, 4 (2018).

14. Capecchi, B. et al. *Neisseria meningitidis* NadA is a new invasin which promotes bacterial adhesion to and penetration into human epithelial cells. Mol Microbiol 55, 687–698 (2005).

15. Buckwalter, C.M., Currie, E.G., Tsang, R.S.W. & Gray-Owen, S.D. Discordant Effects of Licensed Meningococcal Serogroup B Vaccination on Invasive Disease and Nasal Colonization in a Humanized Mouse Model. J Infect Dis 215, 1590–1598 (2017).

16. Comanducci, M. et al. NadA diversity and carriage in *Neisseria meningitidis*. Infect Immun 72, 4217–4223 (2004).

17. Gbesemete, D. et al. Protocol for a controlled human infection with genetically modified *Neisseria lactamica* expressing the meningococcal vaccine antigen NadA: a potent new technique for experimental medicine. BMJ Open 9, e026544 (2019).

18. van der Ende, A., Hopman, C.T. & Dankert, J. Multiple mechanisms of phase variation of PorA in *Neisseria meningitidis*. Infect Immun 68, 6685–6690 (2000).

19. Fagnocchi, L., Pigozzi, E., Scarlato, V. & Delany, I. In the NadR regulon, adhesins and diverse meningococcal functions are regulated in response to signals in human saliva. J Bacteriol 194, 460–474 (2012).

20. Alamro, M. et al. Phase variation mediates reductions in expression of surface proteins during persistent meningococcal carriage. Infect Immun 82, 2472–2484 (2014).

21. Bidmos, F.A. et al. Persistence, replacement, and rapid clonal expansion of meningococcal carriage isolates in a 2008 university student cohort. J Clin Microbiol 49, 506–512 (2011).

22. Masignani, V., Pizza, M. & Moxon, E.R. The Development of a Vaccine Against Meningococcus B Using Reverse Vaccinology. Front Immunol 10, 751 (2019).

23. Gorringe, A.R. et al. Phase I safety and immunogenicity study of a candidate meningococcal disease vaccine based on *Neisseria lactamica* outer membrane vesicles. Clin Vaccine Immunol 16, 1113–1120 (2009).

24. Harrison, O.B. et al. Description and nomenclature of *Neisseria meningitidis* capsule locus. Emerg Infect Dis 19, 566–573 (2013).

25. Frye, S.A., Nilsen, M., Tonjum, T. & Ambur, O.H. Dialects of the DNA uptake sequence in Neisseriaceae. PLoS Genet 9, e1003458 (2013).

26. Arreaza, L., Salcedo, C., Alcala, B. & Vazquez, J.A. What about antibiotic resistance in *Neisseria lactamica*? J Antimicrob Chemother 49, 545–547 (2002).

27. Fink, K. Origin and Function of Circulating Plasmablasts during Acute Viral Infections. Front Immunol 3, 78 (2012).

28. Crotty, S., Aubert, R.D., Glidewell, J. & Ahmed, R. Tracking human antigen-specific memory B cells: a sensitive and generalized ELISPOT system. J Immunol Methods 286, 111–122 (2004).

29. Weisel, F. & Shlomchik, M. Memory B Cells of Mice and Humans. Annu Rev Immunol 35, 255–284 (2017).

30. Pennington, S.H. et al. Polysaccharide-Specific Memory B Cells Predict Protection against Experimental Human Pneumococcal Carriage. Am J Respir Crit Care Med 194, 1523–1531 (2016).

31. Jong, W.S. et al. An autotransporter display platform for the development of multivalent recombinant bacterial vector vaccines. Microb Cell Fact 13, 162 (2014).

32. Smith, A. et al. A Live Attenuated Influenza Vaccine Elicits Enhanced Heterologous Protection When the Internal Genes of the Vaccine Are Matched to Those of the Challenge Virus. J Virol 94 (2020).

33. Osterholm, M.T., Kelley, N.S., Sommer, A. & Belongia, E.A. Efficacy and effectiveness of influenza vaccines: a systematic review and meta-analysis. Lancet Infect Dis 12, 36–44 (2012).

34. Thorstensson, R. et al. A phase I clinical study of a live attenuated *Bordetella pertussis* vaccine--BPZE1; a single centre, double-blind, placebo-controlled, dose-escalating study of BPZE1 given intranasally to healthy adult male volunteers. PLoS One 9, e83449 (2014).

35. Solans, L. et al. IL-17-dependent SIgA-mediated protection against nasal *Bordetella pertussis* infection by live attenuated BPZE1 vaccine. Mucosal Immunol 11, 1753–1762 (2018).

36. Wilk, M.M. et al. Immunization with whole cell but not acellular pertussis vaccines primes CD4 TRM cells that sustain protective immunity against nasal colonization with *Bordetella pertussis*. Emerg Microbes Infect 8, 169–185 (2019).

37. Park, C.O. & Kupper, T.S. The emerging role of resident memory T cells in protective immunity and inflammatory disease. Nat Med 21, 688–697 (2015).

38. Perdomo, C. et al. Mucosal BCG Vaccination Induces Protective Lung-Resident Memory T Cell Populations against Tuberculosis. mBio 7 (2016).

39. Wang, H., Hoffman, C., Yang, X., Clapp, B. & Pascual, D.W. Targeting resident memory T cell immunity culminates in pulmonary and systemic protection against Brucella infection. PLoS Pathog 16, e1008176 (2020).

40. Flacco, M.E. et al. Immunogenicity and safety of the multicomponent meningococcal B vaccine (4CMenB) in children and adolescents: a systematic review and meta-analysis. Lancet Infect Dis 18, 461–472 (2018).

41. Mukhopadhyay, T.K. et al. Rapid characterization of outer-membrane proteins in *Neisseria lactamica* by SELDI-TOF-MS (surface-enhanced laser desorption ionization-time-of-flight MS) for use in a meningococcal vaccine. Biotechnol Appl Biochem 41, 175–182 (2005).

42. Bertoldi, I. et al. Exploiting chimeric human antibodies to characterize a protective epitope of Neisseria adhesin A, one of the Bexsero vaccine components. FASEB J 30, 93–101 (2016).

43. Gorringe, A.R. et al. Experimental disease models for the assessment of meningococcal vaccines. Vaccine 23, 2214–2217 (2005).

44. Cecchini, P. et al. The soluble recombinant *Neisseria meningitidis* adhesin NadA(Delta351–405) stimulates human monocytes by binding to extracellular Hsp90. PLoS One 6, e25089 (2011).

45. Buisman, A.M., de Rond, C.G., Ozturk, K., Ten Hulscher, H.I. & van Binnendijk, R.S. Long-term presence of memory B-cells specific for different vaccine components. Vaccine 28, 179–186 (2009).

46. Crotty, S. T follicular helper cell differentiation, function, and roles in disease. Immunity 41, 529–542 (2014).

